# Perioperative diffuse optical imaging of blood flow distributions for porcine skin flap viability assessment

**DOI:** 10.64898/2026.02.13.26346288

**Authors:** Samaneh Rabienia Haratbar, Fatemeh Hamedi, Mehrana Mohtasebi, Li Chen, Lesley Wong, Guoqiang Yu, Lei Chen

## Abstract

**Significance:** Mastectomy skin flap necrosis remains a major complication in implant-based breast reconstruction due to inadequate tissue blood flow. Existing diagnostic technologies are limited by shallow depth sensitivity, dye-related risks, contact requirements, and an inability to continuously assess blood flow.

**Aim:** This study aimed to translate a noncontact, dye-free, depth-sensitive speckle contrast diffuse correlation tomography (scDCT) technique to a clinically relevant porcine skin flap model for assessing flap blood flow and viability.

**Approach:** The scDCT system was optimized to image blood flow over seven days in four porcine skin flaps including Sham (SH), Implant (IM), Half Necrosis (HN), and Full Necrosis (FN). Measurements were compared with indocyanine green angiography (ICG-A) as a reference standard.

**Results:** scDCT enabled longitudinal monitoring of flap blood flow, revealing significant flow differences among flap types and over time. FN flaps consistently exhibited the most severe flow impairment, while other flap types showed partial or complete recovery over time, distinguishing nonviable from viable tissue. scDCT measurements demonstrated moderate to strong correlations with ICG-A across time points.

**Conclusions:** The findings support scDCT as a promising perioperative imaging modality for improving flap necrosis risk stratification and surgical decision-making, with future work focused on large-scale validation and clinical translation.

## 1 Introduction

Mastectomy is performed in approximately one-third to one-half of women diagnosed with breast cancer [1, 2]. Breast reconstruction, including expander or implant-based and autologous techniques, is a common procedure following mastectomy and can significantly improve patients’ sexuality, self-esteem, and overall quality of life [3]. However, the success of the reconstruction is often limited by mastectomy skin flap necrosis (MSFN), the most common postoperative complication, affecting up to 40% of patients [4–6]. MSFN is more prevalent in patients with established risk factors, including smoking, diabetes, obesity, large breast size, and prior radiation therapy [7, 8].

MSFN occurs when inadequate blood flow and oxygen delivery fail to sustain skin-flap viability [1, 9–12]. It can manifest as partial-thickness necrosis limited to superficial dermal layers or as full-thickness necrosis involving the entire flap [13]. Because MSFN severity is depth-dependent, full-thickness necrosis substantially increases the risk of wound failure, implant exposure, infection, and reconstructive loss [7, 13]. These complications often delay adjuvant therapy and impose significant aesthetic, psychological, and financial burdens on patients [1, 8]. Consequently, perioperative assessment of deep-tissue hemodynamics is essential for accurately determining MSFN severity and guiding timely clinical intervention.

Traditionally, mastectomy flap viability has been assessed clinically in the intraoperative setting and remains largely subjective, relying on the surgeon’s clinical judgment through qualitative indicators such as skin color, dermal bleeding, capillary refill, and temperature [14, 15]. Multiple studies have demonstrated that these subjective assessments often lack rigor and reliability and are associated with MSFN rates as high as 40% [6]. These limitations underscore the importance of objective assessment and management of tissue ischemia and hypoxia in skin flaps, which are critical for preventing MSFN.

Several noncontact imaging techniques have been explored to identify ischemic/hypoxic skin flaps at risk of necrosis, including laser speckle contrast imaging (LSCI) [16, 17], spatial frequency domain imaging (SFDI) [18, 19], hyperspectral imaging (HSI) [20], and indocyanine green angiography (ICG-A) [21–25]. All of these methods measure tissue blood flow or oxygenation only in the superficial skin layer, lack depth sensitivity, and therefore cannot reliably identify deeper tissue ischemia [16–20]. Among these techniques, ICG-A has demonstrated clinical value by enabling intraoperative identification and excision of ischemic tissue, thereby reducing MSFN rates to 0-14% [14]. However, the technique shows variable accuracy, with a tendency to overpredict necrosis and trigger over tissue excision [14, 21]. Additionally, several challenges limit its widespread clinical use, including allergic reactions, brief imaging windows (∼5 min), lack of capability for continuous monitoring, poor patient tolerance for repeated postoperative measurements, high cost, and variability in both device performance and operator skill [1, 26–28]. To address these limitations, we developed a novel, noncontact, dye-free speckle contrast diffuse correlation tomography (scDCT; U.S. Patent #9861319, 2018) system capable of depth-sensitive imaging of blood flow distributions in skin flaps [29–31]. The scDCT system uses a galvo mirror to deliver long-coherence near-infrared (NIR) point illumination to multiple positions within a selected region of interest (ROI). A high-resolution scientific complementary metal oxide semiconductor (sCMOS) camera functions as a 2D detector array, capturing fluctuations in spatial diffuse speckle patterns caused by red blood cell motion within the underlying tissue volume (i.e., tissue blood flow) [30, 32]. Three-dimensional (3D) flow images were generated by implementing the boundary blood flow index (BFI) measurements onto a volumetric mesh and reconstructing them using a finite-element-based algorithm [29–31]. Early clinical evaluations confirmed the feasibility of using scDCT to intraoperatively obtain 3D blood flow distributions in mastectomy skin flaps and demonstrated good agreement between scDCT-derived flow maps and ICG-A perfusion maps [29–31]. However, clinical studies face key limitations, including restriction to intraoperative monitoring, time-consuming 3D image reconstruction, and the low incidence of MSFN, all of which limit comprehensive evaluation of the technology.

More recently, we expanded the single-wavelength scDCT system, which measures only blood flow, into a multi-wavelength scDCT (MW-scDCT) system capable of simultaneously imaging blood flow and oxygenation distributions [33]. The MW-scDCT system was evaluated in a rat model incorporating four flap conditions: sham, implant, half necrosis, and full necrosis, representing varying levels of tissue viability. MW-scDCT enabled perioperative imaging of tissue blood flow and oxygenation over 7 days, revealing significant differences across flap types and time points. Importantly, integrating both blood flow and oxygenation into a multivariable classification model substantially improved discriminative accuracy. These findings support its potential clinical utility as a perioperative monitoring tool to guide decision-making, reduce flap failure risk, and improve reconstructive outcomes [33].

In the present study, we translated the use of scDCT from the small rat model to a human-size porcine model to more accurately replicate the surgical and physiological conditions encountered in clinical mastectomy and implant-based breast reconstruction. The porcine model is highly clinically relevant due to its human-like skin thickness, layered anatomy, dermal vascular plexus architecture, and comparable perfusion volume [34]. Its large skin surface also allows the creation of multiple human-sized flaps that can undergo controlled surgical manipulations representing a spectrum of pathological conditions leading to various degrees of tissue blood flow disruption.

To support this translation, several technical advancements were introduced in the present study. The scDCT hardware and software were optimized to increase data acquisition speed and prevent camera frame loss during large-area imaging of porcine skin flaps. The number of sources and ROI size were expanded to accommodate imaging larger tissue area while maintaining an appropriate balance between spatial resolution and acquisition time. Larger source-detector (S-D) separations were used to achieve deeper tissue penetration in the thicker porcine skin flaps, which lowered signal-to-noise ratio (SNR); therefore, noise-correction steps were incorporated to restore blood flow contrast. Moreover, to address the long processing times associated with 3D reconstruction, a 2D mapping approach was adopted to significantly accelerate data processing. Collectively, these optimizations establish a robust platform for objective assessment of porcine skin flap viability and support future clinical translation. In addition, a commercial ICG-A system (SPY Elite®, Stryker) was used as a clinical reference standard for comparison.

## 2 Methods

### 2.1 Animal Preparation and Surgical Protocol

All animal procedures were approved by the University of Kentucky (UK) Institutional Animal Care and Use Committee (IACUC). Eight female domestic pigs (25-45 kg, 3 months old, White Yorkshire-Landrace mix) were purchased from Oak Hill Genetics and housed in UK Division of Laboratory Animal Resources (DLAR) facilities with social housing and ad libitum access to food and water. Prior to surgery, animals were fasted for 8 hours, sedated with Telazol (4 mg/kg), xylazine (2.2 mg/kg), and butorphanol (0.2 mg/kg), followed by anesthesia with 2.5-5% isoflurane, with physiological parameters continuously monitored by DLAR veterinarians. After induction of anesthesia, pigs were positioned prone, maintained normothermic using forced-air and circulating warm-water blankets, and prepared for surgery by hair removal and skin sterilization.

Six 10 cm × 10 cm dorsal regions (three per side) were marked per animal; four were used for surgical flap creation, including Sham (SH), Implant (IM), Half Necrosis (HN), and Full Necrosis (FN), and two served as naïve areas (NA1, NA2) for normalization (**Fig. 1(a)**). Flaps were created using sterile scalpel incisions and blunt dissection as previously described [33]. Briefly, SH was a C-shaped flap with lateral perfusion preserved; IM included placement of an 80-cc silicone implant (8 cm × 2.1 cm; Sientra Inc., CA, USA); C-shaped HN by elevating the flap with a midline incision and bipolar coagulation to reduce distal collateral flow; and FN flap was generated by fully detaching the flap and coagulating all edges to eliminate the blood supply and limit revascularization (**Fig. 1(b)**). All flaps were then sutured in place and cleaned for imaging. To reduce inter-animal variability, flap locations were rotated among pigs.

**Fig. 1.**
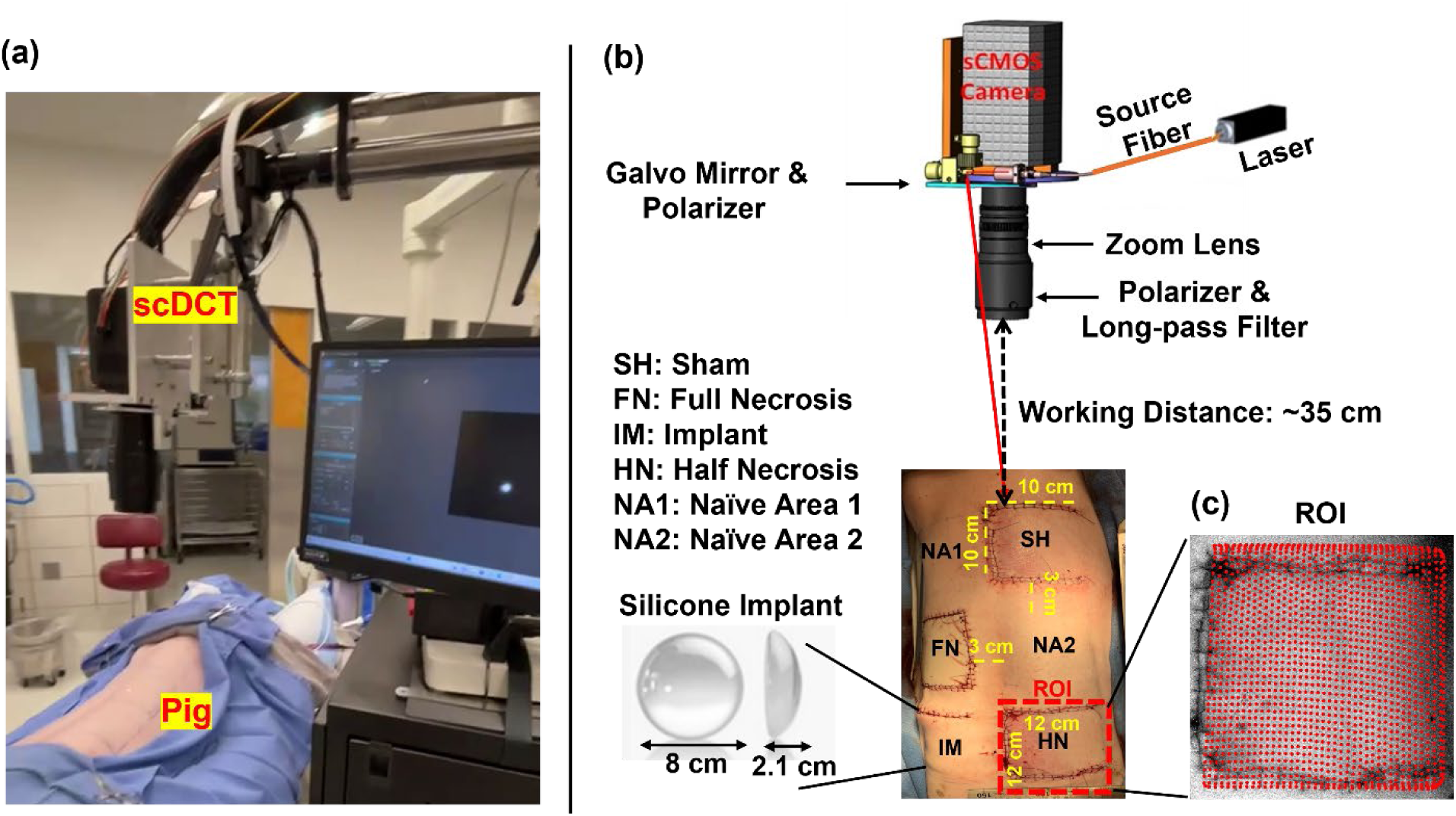
The schematic of optimized scDCT system for imaging of tissue blood flow in porcine skin flaps. **(a**) A photograph of scDCT and experimental setup during data acquisition. (**b**) scDCT principle and instrument design. A galvo mirror was used to deliver point NIR light to multiple source positions. A zoom lens was connected to a sCMOS camera to adjust the size of ROI. A long-pass filter was installed in front of the zoom lens to reduce the impact of ambient light, while a pair of polarizers were added across the source and detection paths to minimize specular reflections from the scanning light sources on the tissue surface. The porcine model includes six areas: four surgical flaps (SH, FN, IM, and HN) and two naïve control areas (NA1 and NA2), each approximately 10 cm × 10 cm with ∼3 cm separation. An 80-cc silicone implant (8 cm × 2.1 cm) was placed beneath the IM flap. (**c**) Source locations (40 × 40) on the selected ROI.

After scDCT and SPY Elite® imaging, anesthesia was discontinued, and animals recovered under DLAR until they were awake, then returned to housing with daily health monitoring. One pig was euthanized early due to an anesthesia-related complication and excluded from analysis. The remaining seven pigs were euthanized at study completion following American Veterinary Medical Association (AVMA) guidelines.

### 2.2 scDCT System Optimization

Details of the scDCT technique for depth-sensitive, and high-density blood flow imaging have been described previously [29, 31, 35]. Briefly, a high-speed galvo mirror delivered coherent NIR light to multiple source positions for boundary data acquisition, while a sCMOS camera captured diffusely reflected light to obtain spatial speckle contrasts within the ROI. Polarizers across the source and detection paths reduced surface reflection, and a zoom lens and a high-performance long-pass filter were used to control the ROI and reduce ambient light interference. Speckle-pattern fluctuations generated by red-blood-cell motion were recorded with the camera exposure time of 5 ms. A custom LabVIEW program, controlled via a data acquisition (DAQ) unit, synchronized the galvo mirror and camera during acquisition. The system sequentially repositioned the galvo mirror and triggered the camera to acquire one frame at each source position sequentially while scanning across the ROI.

The original scDCT system was limited by low temporal resolution (∼0.2 s per source [37]) due to camera constraints in high-speed data acquisition and poor synchronization between the camera and LabVIEW code. Attempts to increase the acquisition rate often resulted in frame loss caused by limitations in data transfer and saving. Storing data temporarily in random access memory (RAM) enabled faster recording but was limited by memory capacity during long acquisition periods, whereas saving directly to the hard drive prevented crashes but resulted in lower temporal resolution.

To address these limitations, several improvements were implemented. The LabVIEW code was revised by adding a trigger-reader from the camera to enhance synchronization. In the updated implementation, once the camera captures one image, a command is immediately sent to the galvo mirror to change position, eliminating the need for a fixed time delay previously used to wait for camera data-saving. In addition, the Hamamatsu camera (ORCA-Flash 4.0, 2048 × 2048 pixels, 30 fps, Hamamatsu, Japan) was replaced with a faster sCMOS camera (Dhyana 400BSI V2, 2048 × 2048 pixels, 74 fps, Tucsen, China), effectively doubling the achievable sampling rate. The upgraded camera supports onboard buffer-based data acquisition, allowing frames to be temporarily stored in internal memory and streamed to disk at high throughput without data loss. This buffered synchronization ensures reliable image acquisition at higher frame rates, eliminating frame loss during high-speed operation. These optimizations improved high-speed synchronization, reduced latency, and increased sampling rate, decreasing the scDCT acquisition time per source from 0.2 s to 0.031 s (6.4-fold reduction).

The previous 2-inch zoom lens (Zoom 7000, Navitar) caused vignetting for large ROIs, resulting in peripheral pixel loss. To overcome this limitation, the lens was replaced with a 2.5-inch zoom lens (TEC-V7X, Computar), enabling vignetting-free imaging over a larger ROI (e.g., ∼ 12 cm × 12 cm) at a working distance of ∼35 cm. A 2.5-inch long-pass filter (LP665; cut-off wavelength: > 665 nm, MidOpt) was incorporated to transmit the 830-nm laser light (DL830-200-SO, CrystaLaser) while eliminating ambient light, together with a compatible custom polarizer (VLR-59.2 mm, Meadowlark Optics) matched to the updated optics. In addition, a simplified LabVIEW-based graphical user interface was developed to streamline device control and improve operational efficiency, which is particularly important for intraoperative use.

### 2.3 scDCT Imaging Procedure

Perioperative scDCT imaging was performed on Days 0, 1, and 7. To ensure consistency across imaging sessions, measurements were acquired using standardized protocols, including uniform animal positioning, working distance, and exposure time, to minimize acquisition variability. At each time-point, the scDCT system was positioned approximately 35 cm above each skin flap to cover a ∼12 cm × 12 cm ROI, encompassing the 10 cm × 10 cm flap and a 1-cm margin of surrounding normal tissue on all sides (**Fig. 1(b)**). To balance temporal and spatial resolution, scDCT data acquired by sequentially scanning 40 × 40 source locations (𝑖 = 1-1600) within the ROI on the porcine back (**Fig. 1(c)**). With a camera exposure time of 5 ms, the total sampling time across all source positions on each skin flap was approximately 50 s (i.e., 1600 sources × 0.031 s per source).

### 2.4 scDCT Data Analysis with Noise Correction

The detailed scDCT data-processing procedures have been described in our previous publications [32, 33]. In this study, an additional noise-correction step was incorporated to improve SNRs, particularly for weaker signals acquired at larger S-D separations [36]. **Fig. 2** illustrates the flowchart for reconstructing tissue blood flow index from scDCT data with noise correction. For each source location (***Si***), the dark offset, determined as the average of 100 dark frames acquired with the laser off (𝐼_𝐷_), is subtracted from the raw intensity image (𝐼) to get the corrected intensity image 𝐼_𝑐_ = 𝐼 − 𝐼_𝐷_. The fundamental speckle contrast (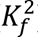) after shot, dark, and quantization noise corrections is given by **Eq. (1)**:

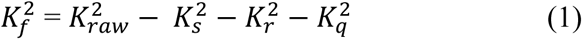

**Fig. 2.**
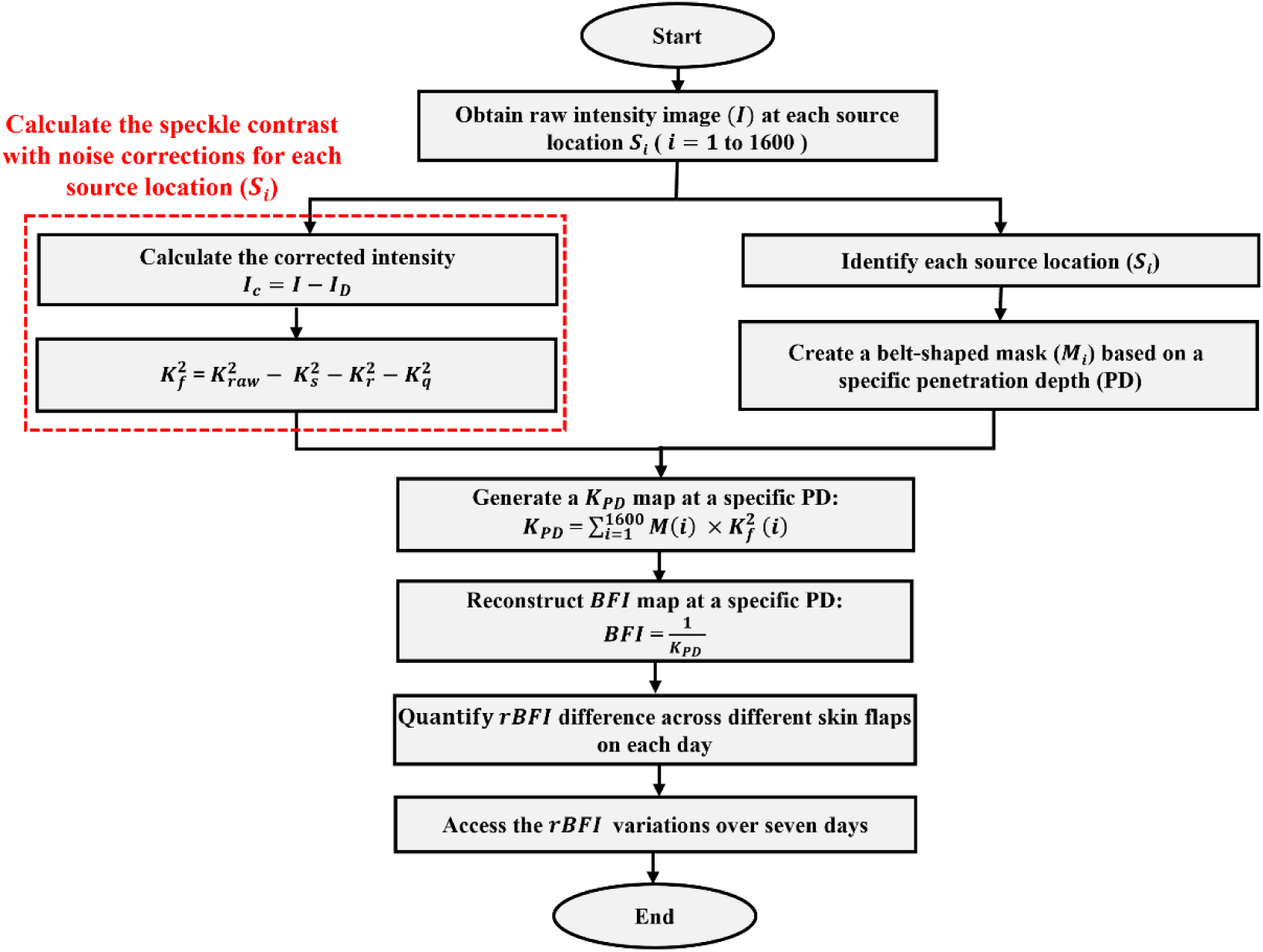
Flowchart illustrating ***rBFI*** quantification over seven days from scDCT data, including noise-correction steps. Red dashed blocks indicate the noise-corrected procedures applied to improve SNR in *BFI* reconstruction.

Here, the raw speckle contrast is defined as 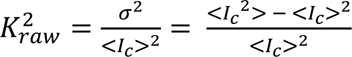, where σ and < 𝐼_𝑐_ > are the standard deviation and mean of intensities within a 7 × 7 pixel window. A sliding-window operation and convolution-based processing were used to generate 2D speckle-contrast maps across the ROI [32, 33].

The speckle contrast arising from photon shot noise is defined as 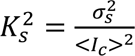 where 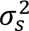 = 𝑔 < 𝐼*_𝑐_* >. Here 𝑔 is the theoretical camera conversion gain, calculated as 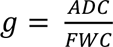, where analog-to-digital converter (ADC) corresponds to 65535 𝐷𝐷𝐷𝐷 for a 16-bit depth, and full-well-capacity (FWC) is 45,000 𝑒⁻, resulting in 𝑔 = 1.45 *DN*/𝑒⁻ . The contrast contribution from camera’s read-out noise is defined as 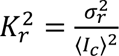, where 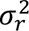 is the temporal variance of a series of 100 dark images. The quantization noise is defined as 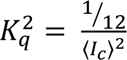, where the quantization-induced bias in variance is assumed to be 1/12 [37, 38]. All noise contributions were subtracted from 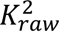 to calculate the noise-corrected 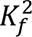 using Eq. (1).

Based on photon diffusion theory, the penetration depth of diffusive light is approximately one-third to one-half of the S-D separation [32]. To generate a special speckle-contrast maps at a specific penetration depth (𝑲***_𝑷D_***), each source location (***Si***) was first identified by converting the intensity image to a binary image using Otsu’s thresholding [39], which automatically selected the optimal threshold to separate bright and dark regions. Connected white pixel groups were then identified, and the largest group was retained as the source position. The center and radius of each source were then determined, and this procedure was repeated for all source positions. A binary belt-shaped mask was created around each source center such that the pixels corresponding to the desired S-D separation (corresponding to the specific ***PD***) were assigned a value of “1”, and all other pixels were assigned a value of “0” [32, 33]. This mask was multiplied by the noise corrected 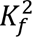 map and summed across all source positions to generate a specific ***K_PD_*** map. To minimize artifacts from overlapping mask regions, the summed maps were normalized by the inverse of the mask-summation image [32].

Given that the mean thickness of porcine skin flaps was approximately 5 mm, S-D separations ranging from 7 to 14 mm were used to achieve an appropriate ***PD*** for generating *K_PD_*map. The blood flow index (***BF1***) map was then generated by calculating ***BF1*** = 𝟏⁄***K_PD_*** Relative ***BF1*** (***rBF1***) values were calculated by normalizing the ***BF1*** map to a normal adjacent area. This normalization was performed on each day, enabling longitudinal comparison of ***rBF1*** changes across flap types and time.

### 2.5 ICG-A (SPY Elite®) Imaging Procedure

ICG-A imaging (SPY Elite®) was performed on Day 0 after the surgery and on Day 7 prior to euthanasia following scDCT measurements. Because of an equipment malfunction, SPY imaging was performed in six of the seven pigs. The SPY system utilized an 805-nm laser to excite ICG fluorescence and a NIR-sensitive camera to detect the emitted fluorescence signals and record real-time video. Animals received an intravenous bolus injection of 2 mL of medical grade, FDA approved ICG solution, prepared by dissolving 25 mg of ICG (Diagnostic Green LLC, MI, USA) in 10 mL of sterile saline. The SPY machine was used to visualize the vascular network and assess blood perfusion within the skin flaps between 3 to 5 minutes after ICG injection, as a reference standard for validating the scDCT measurements.

### 2.6 ICG-A (SPY Elite®) Data Processing

SPY angiography videos were exported as MPEG-4 files and analyzed post-acquisition using a custom MATLAB pipeline. For each flap type, a time window with clearly visible fluorescence was selected. Following the standardized SPY data analysis protocol, a flap-shaped ROI was manually defined, and an adjacent ROI in visually normal tissue was selected for each flap to serve as a local reference for normalization, consistent with the scDCT normalization method. Selected video segments were processed frame by frame and converted to grayscale, and the mean pixel intensity within each ROI was calculated. Similar to scDCT normalization, fluorescence intensity for each flap was normalized to the corresponding adjacent normal tissue to obtain relative fluorescence intensity. Normalized values were then averaged across all relevant frames to yield a single mean fluorescence metric for each flap on Day 0 and Day 7, enabling direct comparison among flap types and over time.

### 2.7 Statistical Data Analysis

SPSS software (Version 29) was used for statistical analysis in animal studies. The repeated measures analysis of variance (ANOVA) was used to evaluate differences in blood flow variations among different days and different flaps. Mauchly’s test of sphericity was first conducted to assess the assumption of sphericity. If Mauchly’s test was not significant, the sphericity-assumed test was used to calculate the overall p-value; otherwise, the Greenhouse-Geisser test was applied. Further, if the overall p-value was significant, post hoc pairwise comparisons were performed to identify the pairwise comparisons with significant differences. The Pearson correlation was calculated to quantify the correlation between scDCT and SPY measurements. A p-value < 0.05 is considered significant for all statistical analysis.

## 3 Results

### 3.1 Clinical Assessment of Skin Flap Viability

**Fig. 3(a)** and **Fig. 4(a)** present the seven-day clinical outcomes for two representative pigs (Pig #1 and Pig #6). The SH and IM flaps showed postoperative recovery, with restoration of normal skin color and texture over the 7-day period. The HN flaps exhibited early darkening in the distal region through Day 1, followed by partial visual improvement over time. In contrast, FN flaps darkened early after surgery and developed a firm texture with no substantial signs of recovery, consistent with complete tissue necrosis.

**Fig. 3.**
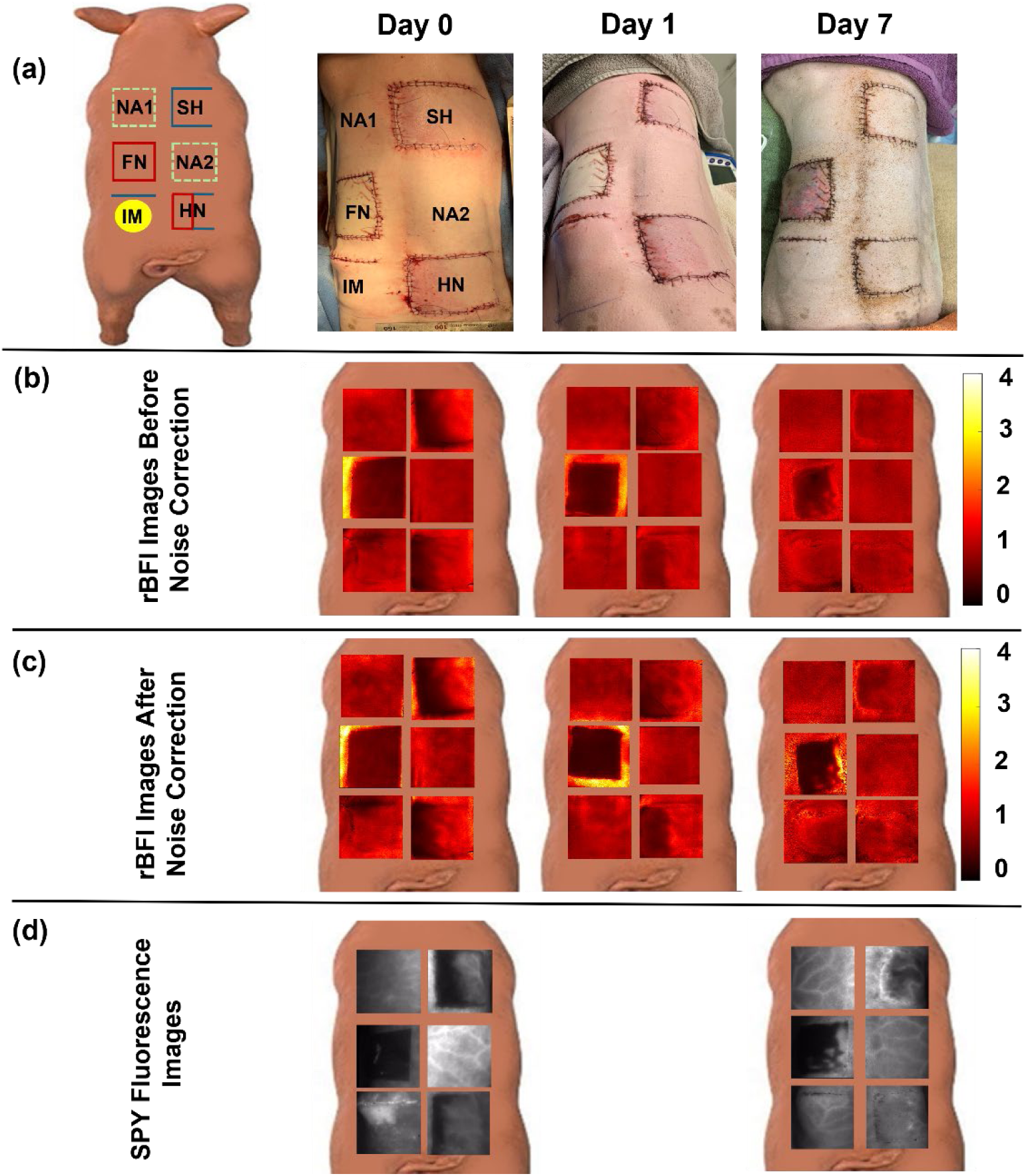
scDCT and SPY imaging of porcine skip flaps in a representative animal (Pig #1). **(a)** Schematic and color photos of four skin flaps (SH, IM, HN, FN) and two naïve control areas (NA1 and NA2) on Days 0, 1, and 7. **(b)** scDCT *rBFI* images acquired on Days 0, 1, 7 before noise correction. **(c)** scDCT *rBFI* images acquired on Days 0, 1, and 7 after noise correction. (**d**) SPY fluorescence images of skin flaps on Days 0 and 7.

**Fig. 4.**
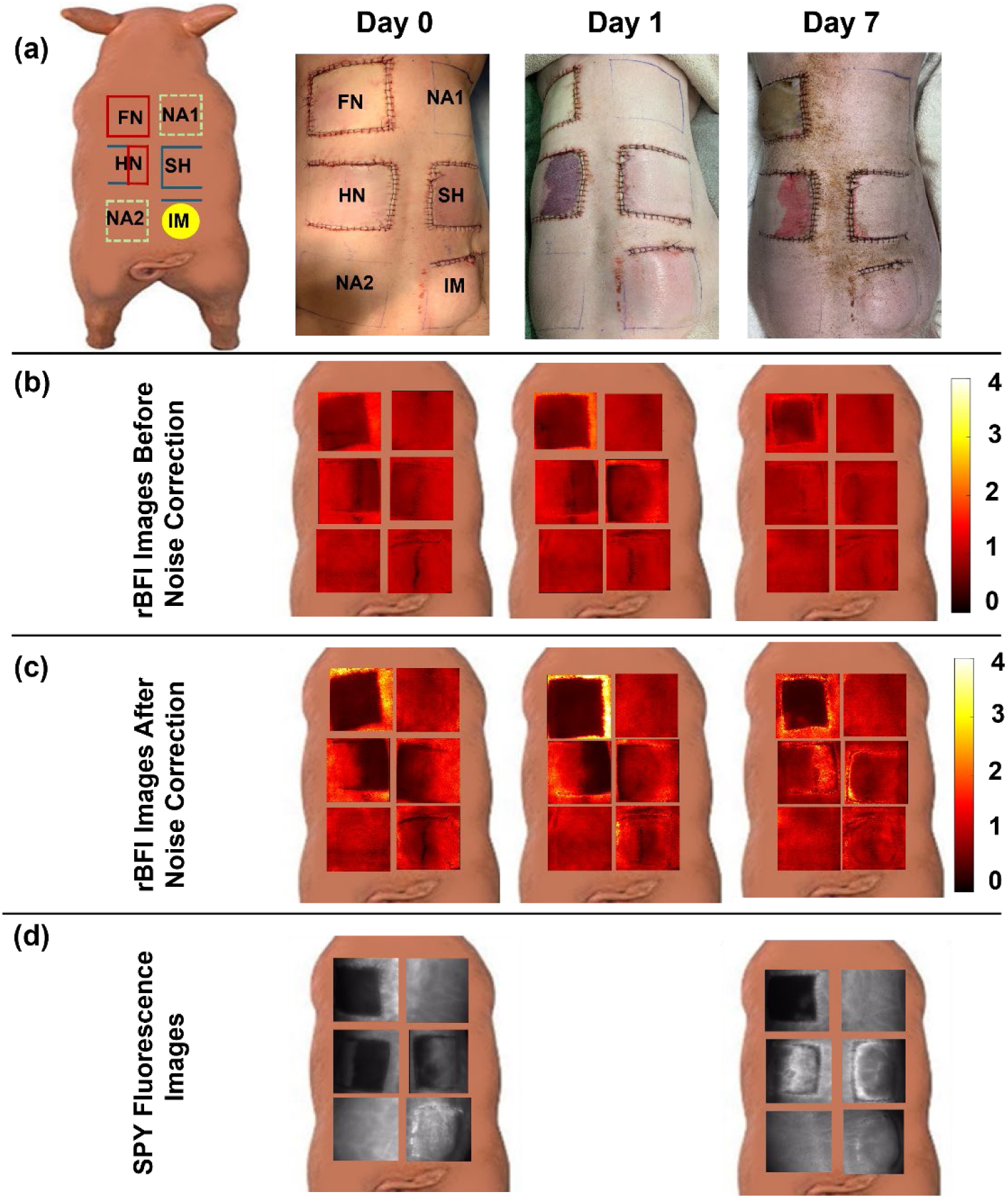
scDCT and SPY imaging of porcine skip flaps for a representative animal (Pig #6). **(a)** Schematic and color photos of four skin flaps (SH, IM, HN, FN) and two naïve control areas (NA1 and NA2) on Days 0, 1, and 7. **(b)** scDCT *rBFI* images on Days 0, 1, 7 before noise correction. **(c)** scDCT *rBFI* images on Days 0, 1, and 7 after noise correction. (**d**) SPY fluorescence images of skin flaps on Days 0 and 7.

### 3.2 Comparable scDCT and SPY Imaging Results in Porcine Skin Flaps

**Fig. 3(b)-3(c)** and **Fig. 4(b)-4(c)** show scDCT measurement results before and after noise correction over the seven-day period for two representative pigs (Pig #1 and Pig #6). The results show distinct temporal blood flow patterns across flap types, and noise correction improved flap differentiation while preserving the overall ***rBF1*** trends observed before correction. On Day 0, immediately after surgery, differences in ***rBF1*** were observed among the flap types, reflecting variability in their initial blood flow responses to surgical manipulation. On Day 1, the FN flaps showed a pronounced reduction in ***rBF1*** compared with the other flaps. By day 7, the SH, IM, and HN flaps in both pigs demonstrated either stable ***rBF1*** values or gradual increases, indicating progressive recovery of blood flow. In contrast, FN flaps displayed persistently low ***rBF1***, with only a slight increase at the edges, consistent with severe ischemia and poor healing potential.

**Fig. 3(d)** and **Fig. 4(d)** show SPY imaging on Day 0 and Day 7, providing complementary perfusion assessment. Bright fluorescence indicates well-vascularized tissue, whereas dark fluorescence reflects ischemia. On Day 0, reduced fluorescence was observed in SH, HN, and FN flaps, indicating impaired perfusion immediately after surgery. By Day 7, fluorescence increased in SH, IM, and HN flaps, suggesting perfusion recovery, while FN flaps remained low or absent at both time points. Overall, SPY trends were consistent with scDCT, supporting distinct vascular responses across flap types and time points.

### 3.3 Skin Flap Differentiation By scDCT and SPY Elite®

**Fig. 5(a)** and **Fig. 5(b)** present group-averaged scDCT results across the different skin flap types over the seven-day period, before and after noise correction. Noise correction generally enhanced flap differentiation and strengthened statistical significance across multiple comparisons while preserving the overall ***rBFI*** trends observed before correction. Accordingly, the following discussion focuses on post-noise-correction results shown in **Fig. 5(b)**.

**Fig. 5.**
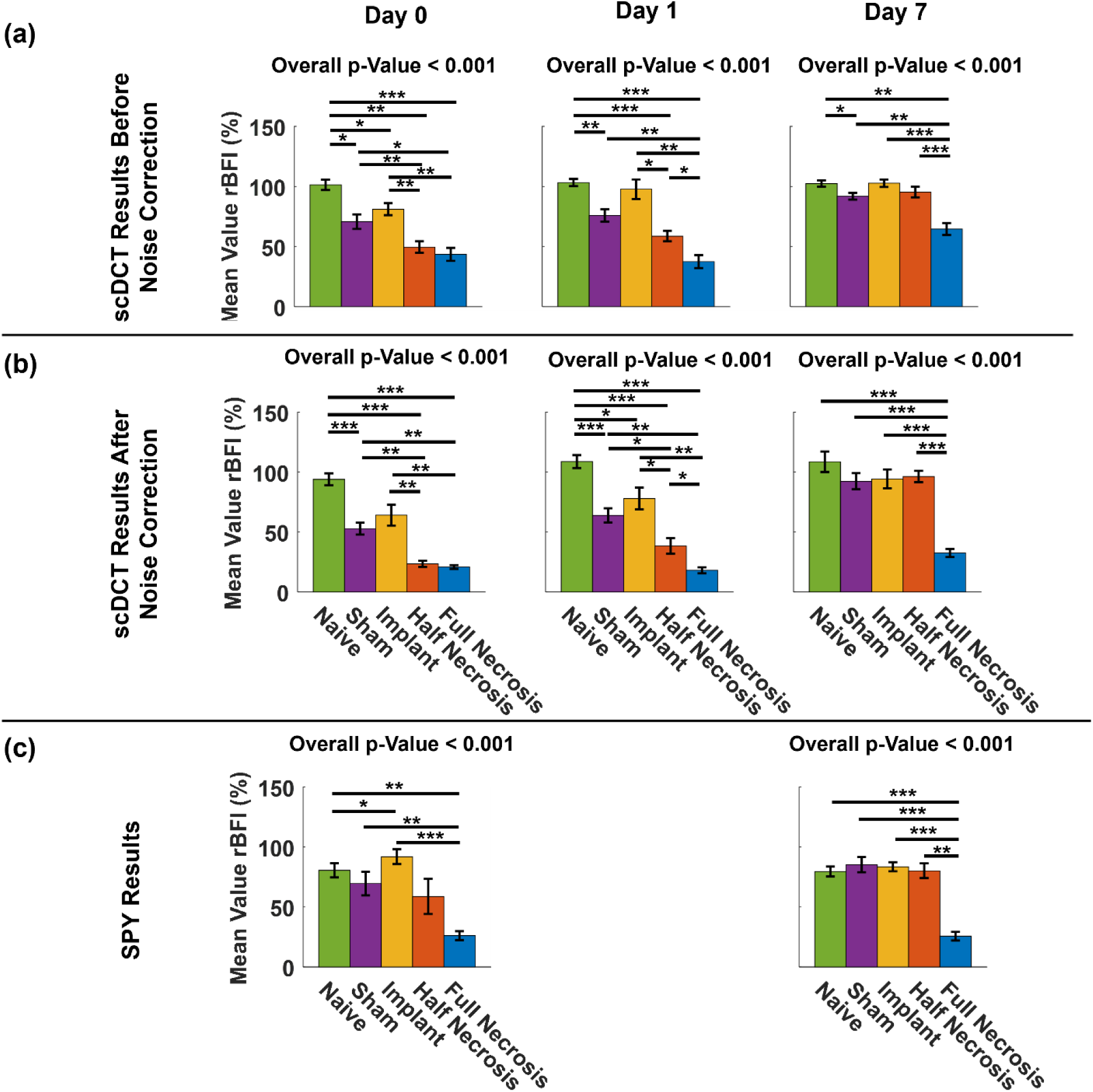
Comparison of group-averaged scDCT *rBFI* results (means ± standard error) before and after noise correction and SPY perfusion results across skin flap types over time. **(a)** scDCT results before noise correction on Days 0, 1, and 7 (n = 7). **(b)** scDCT results after noise correction on Days 0, 1, and 7 (n = 7). **(c)** SPY perfusion results on Days 0 and 7 (n = 6). * p-value < 0.05, ** p-value < 0.01, and *** p-value < 0.001.

On Day 0, NA showed the highest mean ***rBFI*** values, while SH and IM flaps exhibited moderately reduced ***rBFI***. In contrast, HN and FN flaps demonstrated lower ***rBFI*** compared with NA, SH, and IM flaps. Significant differences in ***rBFI*** were observed across different flap types with the overall p-value of < 0.001. Pairwise comparisons showed that HN and FN flaps had significantly lower ***rBF1*** values compared with NA, IM, and SH flaps.

On Day 1, significant differences in ***rBFI*** were observed across flap types with the overall p-value of < 0.001. NA again showed the highest mean ***rBF1*** values, whereas SH and IM flaps exhibited moderately reduced ***rBFI***. HN and FN flaps continued to demonstrate lower ***rBFI*** compared with NA, SH, and IM flaps. Pairwise comparisons showed that HN and FN flaps had significantly lower ***rBFI*** values than all other flap types, with FN exhibiting the lowest values and HN significantly lower than NA and IM. In addition, NA, SH, IM, and HN flaps exhibited ***rBFI*** values comparable to or slightly higher than those on Day 0, indicating early stabilization or partial recovery of blood flow after surgery. In contrast, FN flaps showed a slight decrease in ***rBFI*** from Day 0 to Day 1.

On Day 7, significant differences across flap types persisted with the overall p-value of < 0.001 Pairwise comparisons showed that FN flaps consistently had significantly lower ***rBFI*** than all other flap types, while differences among NA, SH, IM, and HN diminished by Day 7. In addition, all flap types showed ***rBFI*** recovery over time. SH, IM, and HN flaps exhibited increased and comparable ***rBFI*** values by Day 7, while FN flaps also increased from Day 0 and Day 1 but remained the lowest across the seven-day period, indicating persistent hypoperfusion. NA flaps maintained stable perfusion throughout.

**Fig. 5(c)** shows group-averaged SPY results across flap types on Day 0 and Day 7. On Day 0, significant differences in fluorescence intensity were observed across flap types with the overall p-value of < 0.001. Pairwise comparisons showed that FN flaps had significantly lower fluorescence intensity than NA, SH, IM, and HN flaps, indicating severely reduced blood perfusion. NA and SH flaps exhibited moderate fluorescence intensity, consistent with normal or less impaired blood flow immediately after surgery. Notably, IM flaps showed higher fluorescence intensity than NA flaps, likely due to fluorescence dye leakage from disrupted vasculature under increased tension, resulting in localized bright fluorescence and an elevated overall signal.

On Day 7, significant differences in fluorescence intensity were observed across flap types with the overall p-value of < 0.001. Pairwise comparisons showed that FN flaps remained significantly lower than all other flap types, with minimal change over time. HN flaps exhibited a modest, increase from Day 0 to Day 7, while SH, IM, and HN flaps were comparable to NA flaps, indicating restored perfusion and normal tissue viability.

Overall, both scDCT and SPY imaging enabled robust differentiation of skin flap types at early and late postoperative time points, with FN flaps consistently exhibiting the lowest blood flow and perfusion. Both modalities also captured temporal blood flow and perfusion changes, demonstrating recovery across flap types and highlighting their complementary utility for monitoring postoperative vascular dynamics.

### 3.4 Correlation Between scDCT and SPY Measurments

**Fig. 6(a)** and **Fig. 6(b)** show Pearson correlations between scDCT and SPY measurements across all flap types on Day 0 and Day 7, respectively. On Day 0, a moderate and statistically significant correlation was observed between the two measurements (R = 0.64, p < 0.001), although substantial variability was present across flap types. FN flaps clustered at the lower range of both scDCT and SPY measurements, reflecting consistently reduced blood flow and perfusion associated with severe vascular compromise and tissue nonviability.

**Fig. 6.**
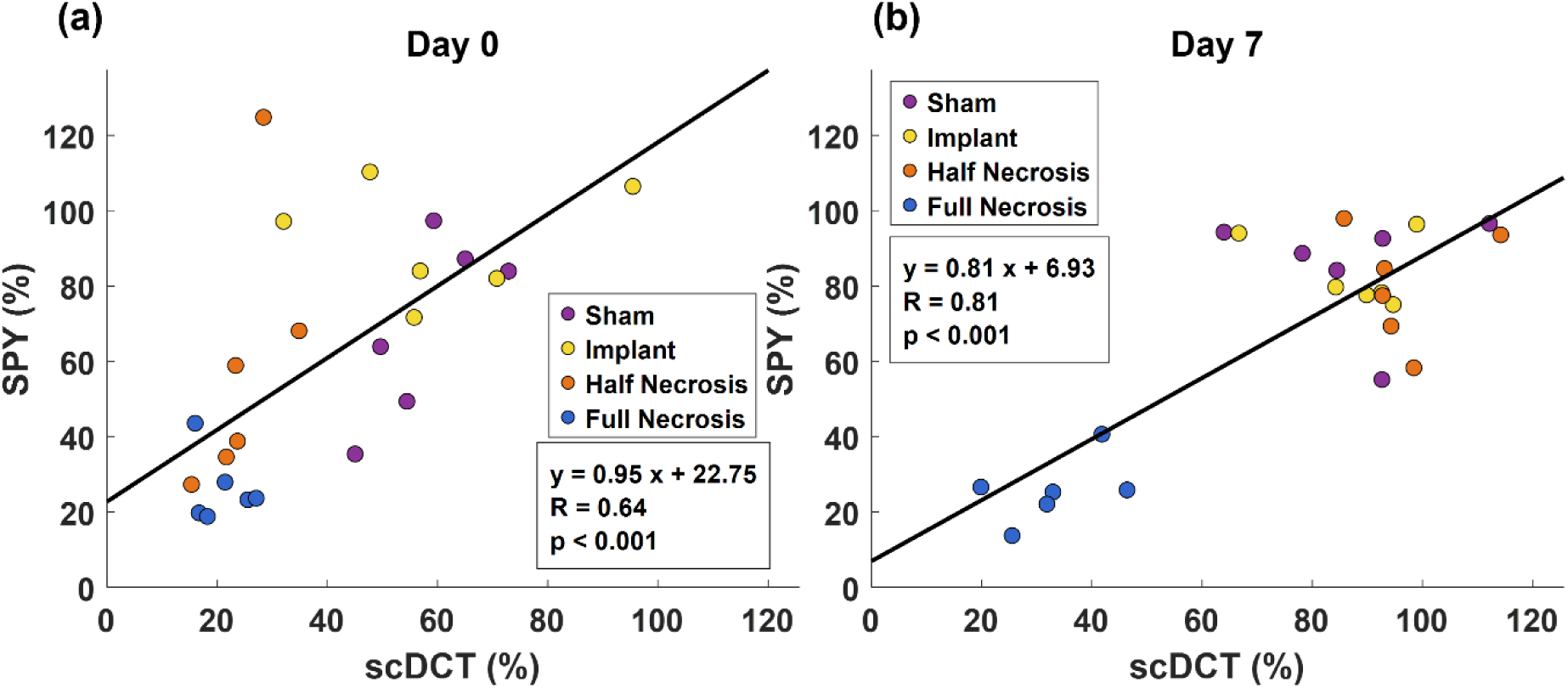
Pearson correlations between scDCT and SPY measurements on Day 0 and Day 7. **(a**) A significant correlation between two measurements was observed on Day 0, although substantial variability was present across flap types. (**b**) A stronger correlation was observed on Day 7, with FN flaps clustering at low values and other flap types shifting to higher values, indicating recovery of blood flow and perfusion.

By Day 7, a strong and statistically significant correlation was observed between the two measurements (*R* = 0.81, p < 0.001). FN flaps again clustered at the lower end of both metrics, consistent with persistently reduced blood flow and perfusion, while the other flap types shifted to higher values, consistent with recovery. Consequently, discrimination between FN and other flap types became more pronounced.

Overall, moderate to strong correlations between scDCT and SPY imaging were observed across flap types and time points, indicating good agreement and consistent assessment of blood flow and perfusion by the two methods.

## 4 Discussion

A major complication following mastectomy and implant-based breast reconstruction is MSFN, which primarily results from inadequate tissue perfusion and oxygen delivery [1, 11, 12, 40, 41].

Perioperative assessment of skin flap viability is therefore critical for identifying tissue at risk for MSFN and guiding timely surgical intervention. In this study, an innovative, dye-free scDCT system previously developed in our laboratory [29, 30, 33] was translated to a clinically relevant porcine skin flap model to enable perioperative depth-sensitive imaging of blood flow distributions (**Fig. 1**). Longitudinal imaging of human-size flaps over a seven-day postoperative period revealed distinct temporal ***rBFI*** changes associated with varying degrees of tissue viability, providing quantitative information directly relevant to MSFN development.

This study incorporated several technical advancements to support clinical translation of scDCT for large porcine skin flap imaging. Improved camera-galvo mirror synchronization, a faster sCMOS camera, and buffered high-speed acquisition substantially increased sampling rate and eliminated frame loss, reducing per-source acquisition time from 0.2 to 0.031 s. Expanded field-of-view optics further enabled vignetting-free imaging over human-size skin flaps. Together, these optimizations improved acquisition efficiency, data reliability, and operational performance while preserving sensitivity to blood flow changes critical for intraoperative use.

Noise-correction procedures (**Fig. 2**) substantially improved SNR and enhanced discrimination between flap types, particularly at larger S-D separations needed for deeper penetration in thick porcine skin flaps. Noise correction strengthened statistical separation between viable and nonviable flaps while preserving temporal blood flow dynamics, underscoring its importance for reliable depth-sensitive imaging in large, heterogeneous tissues.

scDCT enabled longitudinal assessment of blood flow changes in porcine skin flaps, revealing significant differences in ***rBFI*** across flap types and time points (**Fig. 3** and **Fig. 4**). FN flaps exhibited the most severe impairment, with persistently reduced ***rBFI*** throughout the seven-day monitoring period, indicative of prolonged hypoperfusion. In contrast, SH, IM, and HN flaps showed partial or complete blood-flow recovery. Notably, scDCT detected significantly reduced ***rBFI*** immediately after surgery in FN and HN flaps, highlighting its potential for early identification of tissue at high risk for necrosis before irreversible damage occurs.

Group-level analyses verified scDCT sensitivity to blood flow differences across flap types and time points (**Fig. 5**). FN flaps consistently showed the greatest ***rBFI*** impairment, whereas SH, IM, and HN flaps exhibited partial or complete recovery over time, reflecting distinct perfusion patterns between nonviable and viable tissue. Overall, scDCT effectively captured flap-specific blood flow changes and enabled longitudinal monitoring of tissue ischemia and recovery.

Comparison with ICG-A (SPY Elite®) showed moderate to strong correlations with scDCT (**Fig. 6**), with weaker agreement immediately after surgery, likely due to dye-related effects, including transient leakage that can elevate fluorescence independent of true perfusion [42–44]. This was most evident in implant flaps on Day 0. By Day 7, correlations increased substantially as vascular dynamics stabilized, highlighting ability of both imaging modalities to track physiologically meaningful recovery relevant to MSFN outcomes. However, scDCT offers several advantages over SPY imaging for MSFN risk assessment, including dye-free operation and suitability for repeated perioperative measurements without altering tissue physiology. Furthermore, its sensitivity to deep blood flow changes enables detection of tissue ischemia not evident at the surface, making scDCT a valuable complement to current clinical assessments for improving intraoperative decision-making and longitudinal postoperative monitoring.

Several limitations should be noted. Although the porcine model closely approximates human mastectomy skin flaps in size, thickness, and vascular anatomy, the controlled experimental setting may not fully capture clinical complexity. In addition, scDCT was used for observational blood-flow assessment and was not integrated into real-time perioperative decision-making. These limitations will be addressed in future clinical studies. Surface curvature may also have influenced the measurements; however, this effect was minimal given the relatively smooth flap geometry. Geometry-based correction methods will be explored in future work.

## 5 Conclusions

This study demonstrates the promise of scDCT for noninvasive perioperative imaging of blood flow distribution in reconstructive skin flaps using a porcine model. By capturing early and longitudinal blood-flow changes, scDCT reliably differentiated viable and nonviable flap types in a clinically relevant human-size skin flap model. Unlike ICG-A, scDCT enables repeated, dye-free assessment and provides complementary depth-sensitive blood flow information, with good agreement with clinical outcomes. These results support scDCT as a promising perioperative imaging modality to improve flap necrosis risk stratification and surgical decision-making, with future work focused on validation in larger cohorts and translation to human studies with real-time feedback.

## Code, Data, and Materials

All codes and data supporting the findings of this study are available from the corresponding author upon reasonable request.

## Data Availability

All data supporting the findings of this study are available from the corresponding author upon reasonable request.

## Acknowledgments

We acknowledge financial support from the National Institutes of Health (NIH) under grants R01-EB028792, R01-HD101508, R41-NS122722, R42-CA314592, and R42-MH135825 (G.Y.). The content is solely the responsibility of the authors and does not necessarily represent the official views of the NIH.

